# An online mental health informed physical activity intervention for emergency service workers and their families: A stepped-wedge trial

**DOI:** 10.1101/2022.02.10.22270712

**Authors:** Grace McKeon, Ruth Wells, Zachary Steel, Dusan Hadzi-Pavlovic, Scott Teasdale, Davy Vancampfort, Simon Rosenbaum

## Abstract

**Objective:** Emergency service workers are at risk of experiencing poor mental health due to repeated exposure to potentially traumatic events. Promoting physical activity and diet may help to mitigate some the consequences of emergency work and so this study aimed to evaluate the impact a lifestyle intervention on levels of psychological distress among emergency service workers and their support partners.

**Methods:** We delivered a 10-week physical activity intervention via a private Facebook group facilitated by exercise physiologists, a dietitian and peer-facilitators. Weekly education modules and telehealth calls were delivered, and participants were provided with a physical activity tracking device. A stepped-wedge design was applied to compare levels of psychological distress (Kessler-6) during baseline, to intervention by comparing slopes of change. Secondary pre-post outcomes included mental health symptoms, physical activity, quality of life, social support to exercise, sleep quality and suicidal ideation.

**Results:** N=90 participants (n=47 emergency service workers and n=43 support partners) were recruited in 4 separate cohorts (mean age 42.3(SD=11.5) years, 51% male). Levels of psychological distress did not change significantly during the baseline (control) slope and reduced significantly during the first 6 weeks of intervention (intervention slope 1). The interaction between slopes were significant, b=-0.351, *p* = 0.003, (i.e., the trajectories of change were significantly different) and improvements plateaued until the 4-week follow-up. Retention was high (92%) and improvements in mental health symptoms, minutes of physical activity, sedentary time and quality of life were observed.

**Conclusions:** A physical activity intervention delivered via social media is effective in improving psychological distress among emergency service workers and support partners.

**Trial registration:** Australian New Zealand Clinical Trials Registry (ACTRN): 12619000877189.

## Introduction

There are currently over 115,000 full time and 235,000 volunteer emergency service workers in Australia, including paramedics, firefighters, police officers and state emergency service workers (1). People in these occupations work to ensure public health and safety, and in doing so are regularly exposed to significant occupational stressors and potentially traumatic events including violence, natural disasters and death. Emergency service workers are subsequently at increased risk of developing mental health disorders including depression, anxiety and posttraumatic stress disorder (PTSD) (2). A 2018 national survey of emergency services personnel across Australia found that approximately 25% of people currently employed were experiencing high levels of psychological distress and 10% met diagnostic criteria for PTSD (1), compared to 4% in the general population (3). The rates of mental disorders among former employees were even higher (4), with rates of PTSD estimated to be 23% (1). A consequence of these mental health issues is high rates of suicidal thoughts and behaviours (1). In Australia, one emergency service worker dies by suicide approximately every four weeks (5). Maintaining and promoting the mental health of this workforce is essential, not only for employees but for the broader community who rely on their services.

In addition to trauma exposure, emergency service workers face other occupational risks including shift work and highly stressful work environments. Common consequences associated with shift work include poor sleep (6), poor diet (7) and low levels of physical activity (8). In addition, the emergency service worker’s role typically involves primarily sedentary tasks (e.g., desk work or sitting in a vehicle), with only occasional periods of intense physical exertion (9). Sedentary behaviour and physical inactivity are modifiable behaviours associated with an increased risk of poor mental health (10). These risk factors also lead to poor physical health outcomes including high rates of hypertension and obesity which are commonly seen in emergency service workers (11-13).

There is increasing academic and clinical interest in the role of lifestyle interventions targeting physical activity and nutrition behaviours to simultaneously improve physical and mental health (14, 15). Physical activity and its structured subset, exercise, can improve mood and reduce psychiatric symptoms among people with diagnosed mental disorders as well as subclinical populations experiencing mental health symptoms (16, 17). A study among paramedics found that sleep and physical activity explained a significant level of variance in trauma response (35%), indicating likely protective effects of such behaviours and the role of lifestyle interventions as a potentially feasible first-line mental health strategy (18, 19). Emergency service workers require tailored interventions to help overcome some of the adverse health consequences of their work and targeting lifestyle behaviours may be a less stigmatising and culturally acceptable strategy compared to traditional mental healthcare (20). Despite the known mental health and cardioprotective benefits of exercise, there is limited research investigating the role and ways of engaging emergency service workers in such interventions (19).

The potential role of physical activity interventions may be explained by the Conservation of Resources Theory (COR), a stress theory that describes how individuals seek to obtain, retain, protect, and foster valued resources and minimize any threats of resource loss (21). Examples of resources include time, physical energy, social support and self-efficacy. Threats to resource loss occur when individuals are exposed to challenging work and life demands for an extended period, with no opportunity to gain new resources. Resource loss is strongly related to psychological outcomes so applying the COR theory to populations under extreme stress such as emergency service workers is important (22). Hofboll et al. identified that public health promotion efforts in line with the theory would be most effective when applied early to those at risk of experiencing future loss, since individuals who hold greater resources are more capable of resource gain (23). Therefore, by targeting health behaviours such as physical activity and diet, this resource may subsequently improve self-efficacy and the belief that one has the cognitive, physical and emotional resources to engage in their work (24). Physical activity may also prevent the downward spiral of resource loss by not only attenuating stress-induced physiological responses, but by promoting new energetic resources, as suggested by the theory (25). This has been found in a previous study which showed that exercise and high cardiorespiratory fitness can help police officers to better cope with chronic stress and subsequently take less sick leave (26).

Social connections are another important resource which are often lost when people experience mental health issues including PTSD and depression. This also includes the families of emergency service workers who share the burden and impacts of trauma exposure and workplace stress (27). Enhancing social resources can therefore help buffer the effects of loss-related events and is an important health behaviour change strategy (28). Social resources can be proximal including family members and spouses, and distal including friendships with peers. Therefore, group health promotion interventions and the inclusion of already established social networks may be a useful strategy to maximise adherence.

Over a third of the world’s population (38%) use social media sites such as Facebook, Instagram and WhatsApp (29). Given the rapid growth in the use and availability of social media, it may be a scalable and potentially cost-effective opportunity to deliver health promotion interventions to Australia’s geographically dispersed emergency service workforce (30). Online delivery offers flexibility for emergency workers and families to access the intervention in their own time, and based on findings among other populations, can provide social resources through interactions and support between peers (31).

Strategies to help mitigate the mental and physical health consequences of emergency service work and trauma exposure are needed. Physical activity and diet are modifiable risk factors, yet to date there have been no studies targeting these health behaviours and delivered online among emergency service workers. The aim of this study was therefore to evaluate the efficacy of a mental health informed 10-week physical activity and diet program delivered via Facebook for sedentary emergency service workers and their chosen support partners. The effect of the program on psychological distress (primary outcome), depression, anxiety and stress, physical activity levels, sleep quality, quality of life, social support to exercise and suicidal ideation were assessed.

## Methods

### Study design

This study applied a stepped-wedge cluster design to test the impact of the intervention on emergency service worker and support partner outcomes. This involved the sequential roll out of the intervention to four closed cohort clusters over time. We utilised this design given the expectation that the intervention should not be withheld from participants, based on the extant literature on lifestyle interventions improving physical and mental health outcomes (14, 16). This allowed all participants to have timely access to the lifestyle intervention while maintaining the research strengths associated with a randomised clinical trial. The trial was registered with the Australian and New Zealand Clinical Trails registry (ACTRN12619000877189) and approved by the UNSW Human Research Ethics Committee HC180561. The study is reported in line with the CONSORT 2010 checklist extension for stepped wedge cluster randomised trials (32). The study protocol has been published elsewhere (33) and was informed by pilot work (34).

Data were collected on multiple occasions before the intervention (baseline) to detect whether the intervention had an effect significantly greater than the underlying secular trend during baseline. Consecutive observations were interrupted by the intervention to see if the slope (rate of change over time) or level (mean starting point) of the time-series changed following the intervention. Participants were recruited into successive clusters and the baseline lengths of each cluster were randomised between two weeks and five weeks with treatment introduced at different times for each cluster to control for the effects of time as shown in Figure 1. A random number generator in excel was used to allocate the baseline length of each cluster in random order.

## Participants

Participants were recruited between August 2019 and July 2021 through Behind the Seen, a not-for-profit, ex-service organisation that aims to increase awareness and reduce stigma toward mental health issues faced by emergency service workers and their families (35). The Behind the Seen facilitators posted the study advertisement on their Facebook page. Interested participants contacted the research team who provided information about the study and a link to the online screening questionnaires. The screening questionnaires including the Exercise and Sports Science Australia (ESSA) exercise pre-screening tool, the Physical Activity Vital Sign (PAVS), the Kessler-6 (K6) and the Suicidal Ideation Attributes Scale (SIDAS). A member of the research team then checked the questionnaire responses to determine eligibility based on the following criteria: i) former or current emergency service worker; ii) aged 18–65 years; iii) currently physically inactive based on WHO guidelines and defined as engaging in less than 150 mins of moderate–vigorous physical activity per week; iv) absence of any absolute contraindications to exercise as per the ESSA exercise pre-screening tool; v) English speaking and vi) internet and Facebook access. Participants were excluded if they; i) scored >25 in the K6 (indicative of very high levels of psychological distress) and were not receiving treatment and/or medication had changed in the previous 4 weeks, ii) scored ≥21 in the Suicidal Ideation Attributes Scale (SIDAS) (indicative of a high risk of suicidal behaviour), and if the psychologist in the research team determined them ineligible based on a suicide risk assessment.

Eligible emergency service workers were required to nominate a support partner to participate in the program with them. This was defined as any person with a close personal relationship to the emergency service worker (e.g., spouse, family member, caregiver, or close friend). The support partner was required to be i) aged between 18 years and 65 years; ii) absence of any absolute contraindications to exercise as per the ESSA exercise pre-screening tool; iii) English speaking; and iv) internet and Facebook access. The same exclusion criteria for support partners were used. Monte Carlo simulations conducted in Mplus8 using our pilot data (psychological distress (Kessler-6)) showed that a sample size of n=80 would be sufficient to achieve power of 80% with the *P* value set at 0.05 (34).

## Intervention

Participants were enrolled in a 10-week physical activity program delivered via a private Facebook group in 4 separate clusters. The Facebook group was co-facilitated by exercise physiologists and a dietitian with mental health expertise (36) and peer-facilitators (37).

### Exercise Physiologists

The exercise physiologists provided education and encouraged discussions on different predetermined weekly topics including goal setting, overcoming barriers, reducing sedentary behaviour, and nutrition. The content was informed by the qualitative feedback from our pilot study and based on behaviour change techniques including fostering social support, self-monitoring and shaping knowledge (38). All aspects of the program were codeveloped with the facilitators of the community organization (Behind the Seen), who have lived experience of both working as an emergency service worker, living with PTSD and being a spouse of an emergency worker. The exercise physiologists posted 3-4 times per week and monitored the Facebook group daily from Monday to Friday. The dietitian led one week of the intervention which focused on nutrition. The group aims and rules were pinned to the Facebook group so that they were always accessible. Rules included being respectful of other people’s opinions, zero tolerance of rude or hurtful comments, ensuring confidentiality of participants of the group and no sharing of personally identifiable information of others without their permission. Any harmful content which did not align with the aims of the group would be deleted.

### Peer-facilitators

The peer-facilitators were participants from previous clusters of the program who expressed interest to volunteer to take part in a subsequent cluster to help motivate and support other participants. Peer-facilitators in the first cluster were recruited from our pilot study (34). Participants who expressed interest were ‘graduated’ into the role of peer-facilitator for the subsequent cluster of the program and co-delivered the succeeding program with the exercise physiologists. The peer-facilitators were selected based on their engagement with the program, rather than their physical activity levels or experience. The peers were encouraged to post on the Facebook group, participate in weekly group video calls with the facilitators and self-disclose their journey with participants in the group where they were comfortable doing so. No formal training was provided, however the study exercise-physiologists had regular meetings with the peer-facilitators to check in, discuss any concerns and review the progress of the program.

The exercise physiologists and peer-facilitators co-hosted two weekly group video calls via Zoom which lasted approximately 30 mins. Participants were encouraged to call into a single session per week. The study exercise physiologists led the discussion focused on the weekly topic, however the telehealth calls also provided an opportunity to foster social connection and open discussion between participants, peers and the study facilitators. All participants were given a physical activity tracking device (Fitbit Inspire) and the facilitators set up step count challenges using the Fitbit app which involved competing for the highest overall team step count over 4 weeks between emergency workers versus support partners.

## Outcomes

### Data collection

Data were collected from both the emergency service workers and their support partner via self-report questionnaires. We compared emergency service workers and support partners on key baseline demographics including psychological distress and physical activity levels. Data were collected and managed using REDCap electronic data capture tool hosted and managed by Research Technology Services (UNSW Sydney).

## Multiple baseline data

### Primary outcome

The primary outcome was psychological distress. This was measured using the Kessler-6 (K6) which is a reliable and valid, six-item self-report questionnaire (39). The K6 uses a five-point Likert scale with total scores ranging from 6-30. Total scores of between 6 and 18 are classified as not having a probable serious mental illness and >18 as having a probable serious mental illness (40). Psychometric properties include good internal consistency (α=.83) and acceptable discriminative validity (41). The K6 was assessed every week throughout the intervention including the baseline period.

### Secondary outcomes

#### Physical activity (multiple baseline)

Physical activity levels were assessed weekly using the physical activity vital sign (PAVS) (42). The PAVS assesses weekly levels of moderate to vigorous physical activity and takes less than 1 minute to complete. This was assessed weekly throughout the baseline and intervention phase. All other secondary outcomes were assessed prior to the 10-week intervention, post intervention and at 4-weeks follow up.

#### Physical activity (pre, post, follow up)

The SIMPAQ is a five-item clinical tool designed to assess physical activity among populations at high risk of sedentary behaviour (43). The SIMPAQ was adapted to an online version using REDCap. The SIMPAQ traditionally requires an interviewer to form simple calculations to account for hours left in the day based on previous answers, and our online version uses basic formulas to still allow participants to crosscheck responses based on how many hours they need to account for. Total time per week of walking, moderate–vigorous physical activity (MVPA), and sedentary time were assessed.

#### Depression anxiety and Stress

The Depression Anxiety and Stress Scale (DASS-21) is a 21-item self-report instrument that measures the related negative emotional states of depression, anxiety and stress (44). A total score and 3 separate subscales, each with 7 items, were calculated to identify severity ratings for depression, anxiety, and stress. Higher scores represented more severe symptoms. For the depression domain, scores of 0–4 are considered normal, 5-6 mild, 7-10 moderate, 11-13 severe and >14 extremely severe. For anxiety, 0–3 is considered normal, 4-5 mild, 6-7 moderate, 8-9 severe, and >10 extremely severe. For stress, 0-7 is normal, 8-9 mild, 10-12 moderate, 13-16 severe, and >17 extremely severe.

#### PTSD symptoms

The PTSD Checklist for DSM-5 (PCL-5) is a 20-item self-report measure that assesses symptoms of PTSD. Symptom severity scores range from 0 to 80, with a cut-off score of 33 indicating a provisional diagnosis of PTSD (45). A decrease in scores of >10 points indicates a clinically significant change; >5 points indicates a reliable change (45). The PCL-5 was only completed by the emergency service workers and not their nominated support partners.

#### Sleep quality

The Pittsburgh Sleep Quality Index was used to assess the quality and patterns of sleep in the past month (46). A total of 7 sub scores were calculated to yield a global score ranging from 0 to 21. A sum of 5 or greater indicates poor sleep quality.

#### Quality of life

The AQoL-6D quality of life questionnaire has six separately scored dimensions and provides a summary global ‘utility’ score to describe health related quality of life (47). Dimensions include pain, relationships, independent living, mental health, coping and senses. Total scores range from 20 to 99 with higher numbers representing better quality of life.

#### Social support to exercise

The social support and exercise survey was used as an assessment of the level of support individuals making health behaviour changes (exercise) felt they were receiving from family and friends (48). Two separate scores; one for friends and one for family were calculated whereby higher scores represent greater perceived support (48). It was adapted for the purpose of this study to assess the past month instead of the past three months.

#### Suicidal ideation

The SIDAS was used to assess the presence and severity of suicidal thoughts (49). It is specifically developed for online use and consists of five items, each targeting an attribute of suicidal thoughts: frequency, controllability, closeness to attempt, level of distress and impact on daily functioning. The sum of the five items is the overall score (0-50) and higher scores indicate more severe suicidal ideation. A score of >21 indicates a high risk of suicidal behaviour.

#### Feasibility and Acceptability

Retention was defined as; i) retention in the Facebook group for full duration of the 10-week study, ii) participation in the group by actively viewing posts, iii) not missing greater than two consecutive assessment timepoints. Acceptability was assessed at postintervention using the 14-item feasibility and acceptability questionnaire that has been used previously to measure participant responses to a private Facebook group (50). Responses are answered on a 7-point Likert scale (strongly disagree– strongly agree).

### Statistical analysis

#### Comparing multiple baselines to intervention

To examine the effect of the intervention on the primary outcome (K6), data from each cluster was combined and an interrupted time series design was used to compare the rate of change in K6 on multiple occasions before intervention, to the rate of change during, and following the intervention period. A piecewise latent growth curve model (LGCM) was fitted in Mplus 8 base package using the complex analysis function (51). LGCM extends traditional repeated measures analysis of variance by modelling changes in the mean and the variance of initial status (intercept) and the growth rate (slope) simultaneously. We controlled for baseline DASS-21 scores and position (emergency service worker or support partner) on the intercept. The analysis was clustered by pair (emergency service worker and support partner) to account for non-independence of observations within pairs. We note that the length of the baseline was considerably shorter than the intervention so we used the methodology outlined by Rioux and colleagues (52) to address two key factors in modelling the intervention slope. Firstly, change may not be linear over the duration of the intervention. That is, the slope may change direction or magnitude, so there may need to be additional turning points or ‘knots’ in the slope to appropriately model changes over time. Second, there may also be a change in level at the knot point. That is, the new slope created at the knot point may have a different mean starting point (intercept) than the end of the previous slope. We used the following steps. First, visual inspection of the data was used to identify the location for a knot point. Then separate slopes were modelled for a) baseline; b) intervention slope 1; c) intervention slope 2; d) change in level of knot points. Model constraints were used to test for significant differences between slope estimates between baseline and intervention.

Model fit was evaluated using χ^2^ with a scaling correction factor for MLR, comparative fit index (CFI), Tucker–Lewis index (TLI), root mean-square error of approximation (RMSEA) and standardized root mean-square residual (SRMR). Models are considered good fit if CFI > 0.95, TLI > 0.95, RMSEA < 0.05 and SRMR < 0.05, with a χ^2^ P-value or its scaling correction factor for MLR >0.05. The models were estimated under missing data theory using all available information (53).

#### Pre-post intervention tests

To test for differences between pre and post-intervention in secondary outcomes, we used a multivariate omnibus test in mixed models clustered by pair (emergency service and support partnership) with cohort as a random effect, to test whether, across all 5 dependent variables (depression, anxiety, stress, sleep quality and quality of life) there was a significant effect. This was used to control for type one error. Given a significant multivariate test, individual mixed models were then run for each dependent variable. All analyses for the pre-post tests were conducted on SPSS version 27.

## Results

We recruited n=47 emergency service workers and n=43 support partners across four separate clusters (total N=90). The mean age across all clusters was 42.3(SD=11.5) years. In total, 29/47 (62%) of the emergency service workers and n=17/43 (40%) of the support partners were male. The sample size in each cluster ranged from n=19 to n=28. In total, 37/47 (79%) of the emergency service workers were current serving while 21% were retired or medically discharged. Of the total sample, 61% of emergency service/support partner pairs were spouses, 26% were friends, 9% were family members and 4% other (e.g., carer). There was no signiciant difference in age between emergency servie workers and support partners (*P* = 0.08). The baseline demographics for the 4 clusters are shown in table 1.

**Table 1.**
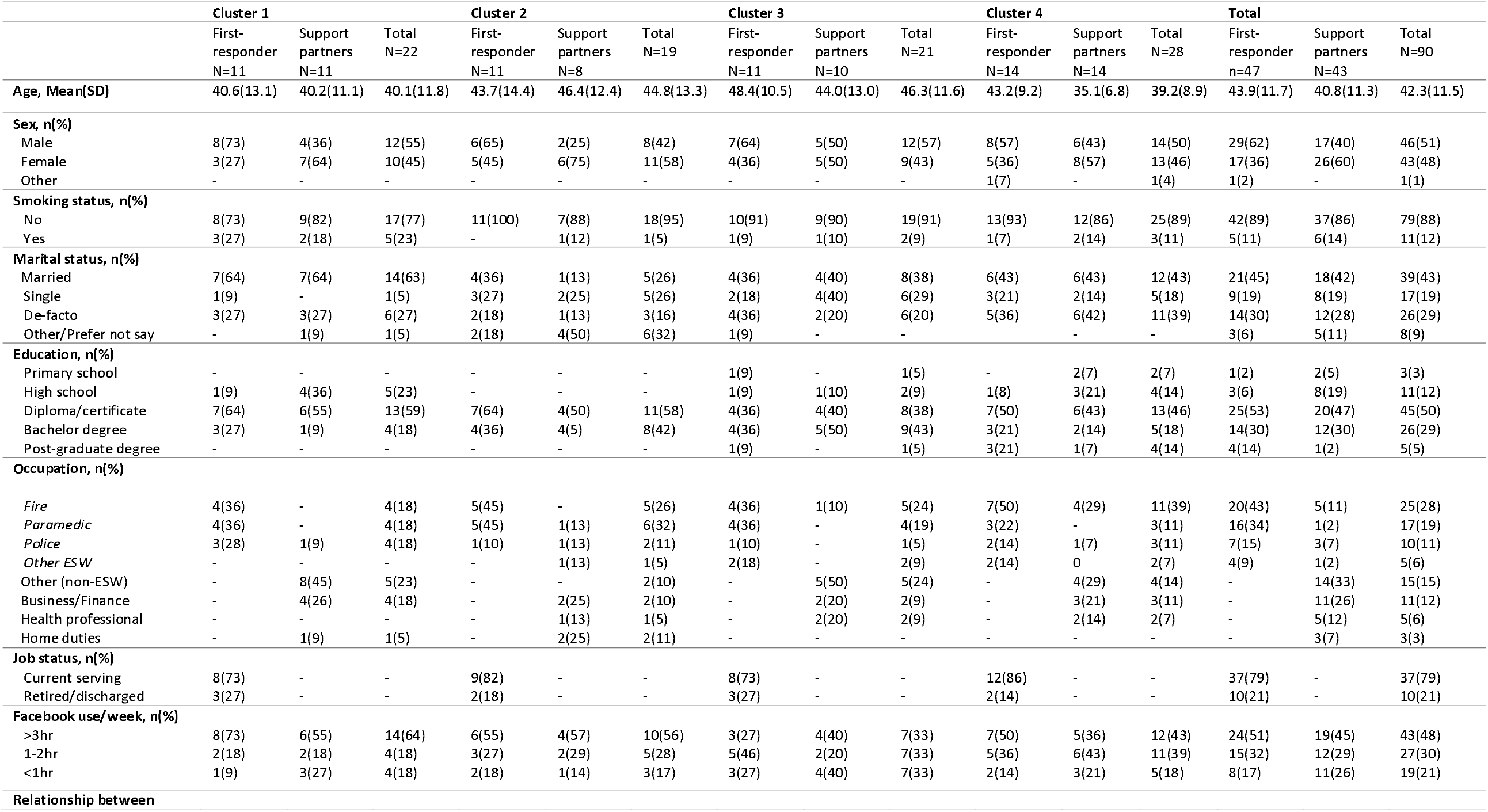
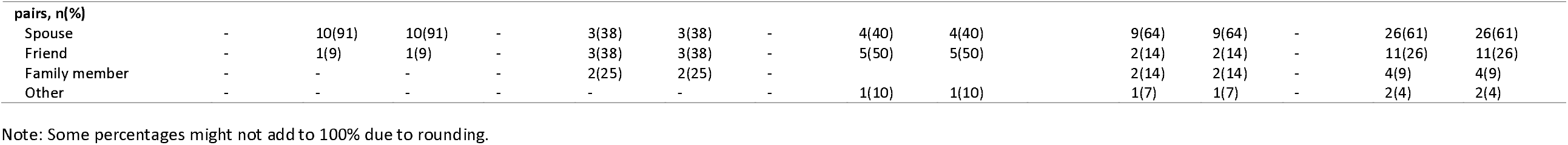
Participant demographics from 4 clusters.

At baseline, n=15 (17%) of the total sample met the cut off score of >18 on the K6, indicating probable serious mental illness. There was no significant difference between the number of emergency service workers vs support partners meeting the cut-off (22% vs 11%, χ^2^ =1.74, N=90, p=0.19). Under half, (n=19, 40%) of the emergency service workers met diagnostic cut off for PTSD at baseline.Mean levels of moderate-to-vigorous activity were 62.3(SD=75.1) mins per week among the emergency service workers and 176.7(SD=261.8) among the support partners. Significantly more support partners versus emergency service workers met the physical activity guidelines at baseline. (37% vs 9%, (χ^2^ =10.31 (1, N=86) =, *P* = 0.001))

Retention was high with 83/90 (92%) completing the intervention. Total dropouts were n=7. Of these, n=2 dropped out during the baseline phase, n=3 before week 5 and n=2 between weeks six and 10. N=53 (59%) completed the one-month follow up. The flow of participants through the trial is shown in Figure 2.

### Psychological distress

Visual inspection of the cluster means revealed a reduction in psychological distress in the first six weeks of the intervention, followed by a floor effect whereby the symptom improvements were maintained at the lower level compared to baseline. That is, there was a change in direction of the intervention slope with a turning or ‘knot’ point approximately halfway through the intervention. We thus modelled the intervention as two slopes, with a knot for a change in direction at week 6 to better represent data. The length of the baseline (four weeks) was more comparable in time course to the two intervention slopes (six weeks each). We therefore had three slopes comprised of four baseline timepoints, followed by six timepoints and another six (including the one month follow up). The first week of the baseline only included cluster 4 (the only cluster with 5 weeks of baseline) so the slope was unduly influenced by this cluster, biasing the slopes. This first week was removed so that only weeks with two or more clusters were modelled.

The baseline slope (slope 1) was not significant, b = -0.09, *P* = 0.379. The Intervention slope 2 (intervention timepoints 1 to 5) showed a significant decrease in psychological distress across the intervention period, b = -0.441, *P* = <0.001. That is, participants decreased by 0.441 of a point on the K6 per week during the first six weeks. The intervention slope 3 (timepoint 11 to 16 (follow-up)) did not decrease significantly, b = -0.009, *P* = 0.883 (i.e., improvements in K6 were maintained). The interaction between baseline and intervention slope 1 was significant, b=-0.351, *P* = 0.003, (i.e., the trajectories of change were significantly different), while slope 2 was not significantly different to baseline b=0.081, *p* = 0.533). That is, participants continually reduced their levels of psychological distress compared to baseline until week six (i.e., improvements were greater in the first part of the intervention) and then improvements plateaued. The model showed mediocre fit (Chi Square = 264.24, *P* < 0.001 RMSEA = 0.097; CFI = 0.918; TLI = 0.913, SRMR 0.114). Clustering cohort revealed that no cluster variables were significant and adding it to the model caused estimation problems, making it unstable. We therefore did not cluster for cluster. The estimated marginal means for K6 scores at each timepoint are shown below in Figure 3. Due to large amounts of variance in reporting of physical activity (see Figure 4 for weekly means), physical activity could be not added to the latent growth curve model.

#### Pre-post tests

The results for all secondary outcomes at baseline and post-intervention are shown in Table 2. The multivariate omnibus test across all variables was significant, i.e., time was significantly associated with the multivariate outcome F (822) = 4.912, *P* < 0.001. Individual mixed models analysis of the secondary outcomes revealed a significant effect of time for anxiety (*P* = 0.02), stress (*P* = 0.003), quality of life (*P* = 0.001), MVPA (*P* = 0.03), walking time (*P* = 0.002), sedentary behaviour (*P* = 0.045), support to exercise from family (*P* = 0.004) and PTSD symptoms (*p* = 0.001). There was no significant effect of time for sleep quality (*P* = 0.07), depression (*P* = 0.15), suicidal ideation (*P* = 0.45) and support to exercise from friends (*P* = 0.19). The effect of position (i.e., emergency service worker versus support partner) was significant across timepoints for depression, stress, quality of life and sleep quality. However, there was no interaction between time and position (i.e., change from pre, post to follow up) between the two groups in any outcome except friends support to exercise.

**Table 2.** Latent growth curve models for psychological distress.

|  | Coefficient | S.E | *t*-score | P value |
| --- | --- | --- | --- | --- |
| **Position on intercept** | 0.658 | 0.553 | 1.190 | 0.234 |
| **DASS-21 on intercept** | 0.277 | 0.030 | 9.148 | **<0.001** |
| **Baseline** | -0.090 | 0.102 | -0.880 | 0.379 |
| **Intervention 1** | -0.441 | 0.062 | -7.098 | **<0.001** |
| **Intervention 2** | -0.009 | 0.064 | -0.147 | 0.883 |
| **Intervention 1 vs baseline** | -0.351 | 0.116 | -3.018 | **0.003** |
| **Intervention 2 vs baseline** | 0.081 | 0.129 | 0.624 | 0.533 |
| **Intervention 1 vs 2** | -0.432 | 0.089 | -4.861 | **<0.001** |

Note: Baseline= slope 1, Intervention 1=slope 2, Intervention 2= slope 3

**Table 3.** Mixed models pre, post and follow up data.

| Variable | n | Baseline-Estimate (SE) | 95% Confidence<br>interval | n | Post -<br>Estimate (SE) | 95% Confidence<br>interval | n | Follow-up<br>Estimate (SE) | 95% Confidence<br>interval | Time |  | Position |  | Time<br>x<br>positi<br>on |  |
| --- | --- | --- | --- | --- | --- | --- | --- | --- | --- | --- | --- | --- | --- | --- | --- |
|  |  |  |  |  |  |  |  |  |  | F test | P value | F test | P value | F test | P value |
| **DASS Depression** |  |  |  |  |  |  |  |  |  |  |  |  |  |  |  |
| Total | 90 | 6.0(0.5) | [4.91, 7.08] | 73 | 5.3(0.6) | [4.20, 6.44] | 50 | 5.3(0.6) | [4.09, 6.56] | 1.9 | 0.15 | 10.02 | **0.002** | 0.53 | 0.59 |
| ESW |  | 7.6(0.8) | [6.12, 9.12] |  | 7.2(0.8) | [5.63, 8.72] |  | 6.8(0.8) | [5.17, 8.48] |  |  |  |  |  |  |
| Partner |  | 4.4(0.8) | [2.90, 5.94] |  | 3.5(0.8) | [1.84, 5.10] |  | 3.8(0.9) | [2.00, 5.65] |  |  |  |  |  |  |
| **DASS Anxiety** |  |  |  |  |  |  |  |  |  |  |  |  |  |  |  |
| Total | 90 | 4.5(0.4) | [3.64, 5.32] | 73 | 3.3(0.5) | [2.82, 4.58] | 50 | 5.3(0.6) | [2.35, 4.32] | 4.1 | **0.02** | 1.39 | 0.24 | 0.46 | 0.63 |
| ESW |  | 5.2(0.6) | [4.01, 6.34] |  | 4.1(0.6) | [2.94, 5.35] |  | 3.6(0.7) | [2.30, 4.94] |  |  |  |  |  |  |
| Partner |  | 3.8(0.6) | [2.57, 5.01] |  | 3.3(0.6) | [1.98, 4.53] |  | 3.1(0.7) | [1.59, 4.52] |  |  |  |  |  |  |
| **DASS Stress** |  |  |  |  |  |  |  |  |  |  |  |  |  |  |  |
| Total | 90 | 4.5(0.4) | [6.76, 8.89] | 73 | 3.7(0.4) | [5.34, 7.57] | 50 | 3.3(0.5) | [4.90, 7.43] | 5.9 | **0.003** | 4.8 | **0.031** | 0.04 | 0.96 |
| ESW |  | 9.0(0.7) | [7.53, 10.47] |  | 7.5(0.8) | [5.99, 9.05] |  | 7.2(0.9) | [5.53, 7.01] |  |  |  |  |  |  |
| Partner |  | 6.7(0.8) | [5.12, 8.19] |  | 5.4(0.8) | [3.77, 7.02] |  | 5.1(1.0) | [3.23, 7.01] |  |  |  |  |  |  |
| **Quality of life** |  |  |  |  |  |  |  |  |  |  |  |  |  |  |  |
| Total | 88 | 75.0(1.5) | [72.14, 77.92] | 72 | 78.5(1.5) | [75.54, 81.52] | 49 | 77.9(1.6) | [74.61, 81.18] | 7.4 | **0.001** | 8.9 | **0.004** | 1.4 | 0.26 |
| ESW |  | 69.73(2.0) | [65.74, 73.73] |  | 74.6(2.1) | [70.48, 78.70] |  | 74.6(2.2) | [70.48, 78.70] |  |  |  |  |  |  |
| Partner |  | 80.4(2.1) | [76.14, 84.56] |  | 82.5(2.2) | [78.13, 86.8] |  | 81.2(2.5) | [76.33, 86.04] |  |  |  |  |  |  |
| **MVPA,**<br>**minutes/week** |  |  |  |  |  |  |  |  |  |  |  |  |  |  |  |
| Total | 89 | 103.7(17.4) | [69.38, 137.94] | 77 | 151.2(18.3) | [114.98, 187.46] | 46 | 135.5(23.4) | [89.37, 181.57] | 3.5 | **0.03** | 2.7 | 0.11 | 2.11 | 0.13 |
| ESW |  | 59.9(24.1) | [12.47, 107.57] |  | 114.72(24.96) | [65.54, 163.90] |  | 142.4(29.0) | [85.18, 199.63] |  |  |  |  |  |  |
| Partner |  | 147.33(24.9) | [97.97, 196.68] |  | 187.72(26.98) | [134.48, 240.96] |  | 128.53(36.7) | [56.24, 200.83] |  |  |  |  |  |  |
| **Sleep quality** |  |  |  |  |  |  |  |  |  |  |  |  |  |  |  |
| Total | 89 | 9.3(0.5) | [8.44, 10.25] | 71 | 8.6(0.5) | [7.67, 9.55] | 44 | 8.8(0.5) | [7.75, 9.89] | 2.7 | 0.07 | 7.772 | **0.006** | 1.09 | 0.34 |
| ESW |  | 10.35(0.6) | [9.10, 11.61] |  | 9.63(0.66) | [8.33, 10.92] |  | 10.42(0.7) | [8.95, 11.90] |  |  |  |  |  |  |
| Partner |  | 8.32(0.66) | [7.02, 9.63] |  | 7.59(0.7) | [6.23, 8.96] |  | 7.22(0.7) | [5.66, 8.77] |  |  |  |  |  |  |
| **Sedentary hours/day** |  |  |  |  |  |  |  |  |  |  |  |  |  |  |  |
| Total | 89 | 8.6(0.4) | [7.85, 9.47] | 72 | 7.7(0.4) | [6.85, 8.61] | 46 | 7.3(0.6) | [6.17, 8.36] | 3.2 | **0.045** | 1.09 | 0.299 | 0.67 | 0.51 |
| ESW |  | 9.37(0.6) | [8.24, 10.50] |  | 8.14(0.6) | [6.94, 9.35] |  | 7.25(0.7) | [5.89, 8.62] |  |  |  |  |  |  |
| Partner |  | 7.95(0.6) | [6.79, 9.12] |  | 7.31(0.6) | [6.04, 8.58] |  | 7.28(0.9) | [5.56, 9.0] |  |  |  |  |  |  |
| **Walking min/day** |  |  |  |  |  |  |  |  |  |  |  |  |  |  |  |
| Total | 89 | 41.6(5.2) | [31.26, 51.95] | 74 | 63.0(5.7) | [51.76, 74.16] | 46 | 64.6(7.3) | [50.15, 78.97] | 6.6 | **0.002** | 0.573 | 0.451 | 2.2 | 0.12 |
| ESW |  | 34.2(7.3) | [19.85, 48.61] |  | 53.6(7.8) | [38.21, 69.10] |  | 71.5(9.0) | [53.80, 89.24] |  |  |  |  |  |  |
| Partner |  | 48.9(7.54) | [34.10, 63.85] |  | 72.3(8.2) | [56.03, 88.54] |  | 57.6(11.5) | [34.88, 80.32] |  |  |  |  |  |  |
| **Social support to exercise (Family)** |  |  |  |  |  |  |  |  |  |  |  |  |  |  |  |
| Total | 69 | 18.4(1.0) | [16.32, 20.45] | 54 | 22.1(1.1) | [19.87, 24.40] | 37 | 19.9(1.4) | [17.24, 22.63] | 5.9 | **0.004** | 0.819 | 0.369 | 0.01 | 0.99 |
| ESW |  | 17.6(1.4) | [14.73, 20.44] |  | 21.4(1.6) | [18.28, 24.55] |  | 19.0(1.8) | [15.43, 22.59] |  |  |  |  |  |  |
| Partner |  | 19.2(1.5) | [16.20, 22.16] |  | 22.9(1.6) | [19.59, 26.12] |  | 20.9(2.0) | [16.84, 24.89] |  |  |  |  |  |  |
| **Social support to exercise (Friend)** |  |  |  |  |  |  |  |  |  |  |  |  |  |  |  |
| Total | 69 | 14.8(0.9) | [13.15, 16.53] | 54 | 15.8(0.9) | [13.97, 17.56] | 37 | 14.5(1.0) | [12.48, 16.59] | 1.7 | 0.19 | 1.496 | 0.225 | 2.71 | **0.07** |
| ESW |  | 14.5(1.2) | [12.17, 16.83] |  | 13.94(1.3) | [11.46, 16.43] |  | 13.8(1.4) | [11.03, 16.57] |  |  |  |  |  |  |
| Partner |  | 15.2(1.2) | [12.74, 17.62] |  | 17.6(1.3) | [15.00, 20.17] |  | 15.3(1.5) | [12.22, 18.32] |  |  |  |  |  |  |
| **SIDAS** |  |  |  |  |  |  |  |  |  |  |  |  |  |  |  |
| Total |  | 11.4(0.6) | [10.18, 12.66] |  | 11.9(0.6) | [10.61, 13.19] |  | 11.2(0.7) | [9.83, 12.78] | 0.8 | 0.45 | 1.372 | 0.245 | 0.658 | 0.52 |
| ESW |  | 12.35(0.8) | [10.61, 14.03] |  | 12.6(0.8) | [10.84, 14.40] |  | 11.36(0.9) | [9.44, 13.36] |  |  |  |  |  |  |
| Partner |  | 10.5(0.8) | [8.73, 12.33] |  | 11.1(0.9) | [9.32, 13.06] |  | 11.1(1.1.0) | [9.01, 13.41] |  |  |  |  |  |  |
| **PCL-5** |  |  |  |  |  |  |  |  |  |  |  |  |  |  |  |
| ESW | 47 | 28.1(2.7) | [22.60, 33.51] | 40 | 24.0 (2.7) | [18.51, 29.53] | 31 | 22.1(2.8) | [16.42, 27.78] | 7.8 | **0.001** | - | - | - | - |

Note: Estimate= estimated marginal means, SE= standard error. Position indicates emergency service worker or support partners. ESW= Emergency Service Worker. Bold font indicates statistical significance. DASS=Depression, Anxiety and Stress Scale, MPVA= moderate to vigorous physical activity, AQoL-6D= Assessment of Quality of Life, SIDAS= Suicidal Ideation Attributes Scale, PCL-5=PTSD checklist for DSM-5.

### Acceptability

The results from the feasibility and acceptability questionnaire are shown in figure 5. There were no adverse events related to the study reported during the conduct of the intervention, nor was there any inappropriate behaviour, online harassment or privacy concerns in the group which needed to be removed by the facilitators.

## Discussion

The results of this study showed that an online physical activity intervention is a feasible, low resource strategy that can help to alleviate some of the common consequences of trauma exposure including psychological distress. Our 10-week intervention resulted in significant reductions in levels of reported psychological distress among emergency service workers and their support partners. Levels of psychological distress did not change significantly during the baseline period, yet reduced significantly during the first six weeks of the intervention, compared to baseline. Improvements then plateaued and were maintained until the end of the intervention and to the one-month follow-up. This finding is consistent with a previous exercise intervention conducted in police officers which found that the greatest improvements in levels of PTSD were made in the first six weeks of the intervention and then maintained until the end of the program at week 12 (54). Therefore, by helping emergency service workers to maintain social and personal resources associated with lifestyle behaviours, this may help to protect their mental health and prevent resource loss spirals, as explained by the COR Theory (21).

In addition to reductions in psychological distress, participation in the intervention was also associated with improved levels of anxiety, stress, quality of life and PTSD symptoms in line with previous studies (55, 56). The active components of the intervention mediating the improvements in mental health symptoms however are not clear. Given the existing exercise and mental health treatment and prevention literature (16, 57), and the target of the intervention, the observed significant changes in physical activity levels and sedentary behaviour are likely contributing factors. While mean weekly physical activity levels could not be added to the latent growth curve model because of the large variability in self-reported data, Figure 4 appears to show a trend of increasing physical activity levels throughout the intervention compared to baseline. Increasing physical activity levels and reducing sedentary behaviour may therefore be a useful strategy for improving mental health in a primarily sub-clinical population at risk of poor mental health. Core components of the intervention included the supervision by exercise physiologists, as recommended to increase adherence among people with poor mental health (58), and the focus on enjoyment and the emphasis on sustainable, incremental lifestyle change tailored to individual preferences, rather than specific exercise prescription (59).

The study also aimed to facilitate social support, which has been found among other populations including Veterans to drive the relationship between increased exercise and improved mental health (60). Investigation into the resource of social support including the dyad relationship (i.e., emergency service and support partner) in moderating health behaviours is warranted. While support from friends to exercise did not change significantly, changes in family support were, which is not surprising given 70% of dyads were spouses or family members rather than friends. Lastly, retention was high (91%) compared to previous social media delivered physical activity and diet intervention studies among the general population (61) and physical activity interventions among people with anxiety disorders (62). This is an important finding given the barriers to other preventative and treatment based mental health initiatives (63).

### Limitations

Several limitations should be considered. Firstly, the online recruitment via social media introduces a bias toward those who are already active online and technologically literate. Secondly, it is difficult to dismantle the intervention components (e.g., Fitbit, Facebook group, telehealth calls), to inform which component is having the greatest effect and inform future initiatives. While we recruited participants with and without a diagnosed mental illness, we excluded people who were experiencing high levels of symptomatology and were not engaged in other mental health support. Since the present study is not mental health treatment it may therefore not be suitable for people experiencing very high levels of psychiatric symptomatology. In addition, the follow up period of one month may not be long enough to detect sustained changes and future research should consider longer follow up period to determine maintenance of behaviour change.

A strength of this study is the stepped-wedge design whereby everyone had access to the intervention, however importantly while the primary outcome (K6) had a control, the pre-post-follow up data did not. Finally, our sample size calculation determined we would need n=80 participants, and so in our protocol we reported that would recruit five clusters to account for drop out (33). Given our low dropout rate (n=7) we terminated recruitment after 4 trials (n=90) since this exceeded the required n=80 participants.

## Conclusion

Emergency service workers are regularly exposed to trauma as part of their work. We therefore need to equip emergency workers with resources to help buffer the effects of stress and trauma exposure. Results add to the small yet increasing body of evidence showing the impact of targeting modifiable lifestyle behaviours, known predictors of mental health, for populations exposed to trauma. Our stepped-wedge evaluation of a 10-week online lifestyle intervention resulted in significant improvements in levels of psychological distress and may be associated with other health outcomes including quality of life among emergency service workers and their support partners. Future research should consider evaluating the mechanistic factors (e.g., levels of physical activity, social support) as mediators of improvements in levels of psychological distress.

## Data Availability

All data produced in the present study are available upon reasonable request to the authors

## Ethical approval

This study was approved by the UNSW Human Research Ethics Committee HC180561.

## Guarantor

GM.

## Contributorship

GM and SR researched the literature and conceived the study. GM, SR, RW, ZS and DV were involved in the protocol development. GM and SR recruited all participants and GM, SR and ST conducted the study. GM, RW, ZS and DH conducted the data analysis. GM wrote the first draft of the manuscript and all authors reviewed and edited the manuscript and approved the final version.

## Acknowledgments

We would like to acknowledge the contribution of Veronique Moseley and Ross Beckley from Behind the Seen to the co-design of this study.

